# Determinants of General Practitioners’ Initiation of Conversations about Childhood Overweight: A mixed-methods study

**DOI:** 10.64898/2026.03.03.26347173

**Authors:** C.C.A. Delhez, M.A. Adriaanse, H.M.M. Vos, R.C. Vos, R.M.J.J. van der Kleij

## Abstract

**Background:** Childhood overweight is a major health concern with long-term consequences. Dutch guidelines recommend that general practitioners (GPs) screen for overweight in children regardless of visit reason, yet GPs infrequently initiate weight-related conversations.

**Objectives:** To explore what determines GPs’ initiation of conversations about childhood overweight, identify the determinants with the highest potential for change, and examine how, in particular, emotional responses and equanimity, relate to GPs’ intention to initiate conversations.

**Methods:** A cross-sectional survey was conducted among Dutch GPs (in training) between March-May 2025. Behavioural determinants, based on the Theoretical Domains Framework, emotional responses, equanimity, and anticipated behaviour and implementation success were assessed. For all determinants, room for improvement (deviation from maximum), relevance (correlation with anticipated behaviour), and the potential for change (combining these two) were calculated using R. Open-ended responses were analysed using content analysis.

**Results:** 57 GPs completed the survey. Most reported adequate skills (66%), knowledge (61%), and motivation (74%); yet only 25% reported high implementation success. Their behaviour is constrained by a lack of habituation, negative outcome expectancies, failing to remember to act, and a lack of social and organizational support. Emotional responses were evident, with 10–15% of respondents reporting high arousal or clearly positive or negative valence. Valence, but not arousal or equanimity, was positively associated with anticipated intention (*r* = 0.45, *p* < 0.001).

**Conclusion:** Supporting routine weight-related conversations requires strategies to strengthen habit formation, reshape outcome expectancies, support memory, address social and organizational factors, and further explore GPs’ emotion regulation.

**Key messages:**

- Although highly motivated, general practitioners infrequently initiate conversations about childhood overweight due to existing practical barriers and barriers related to internal processes.
- Habit formation showed the greatest potential for change, emphasizing the importance of automaticity in initiating conversations.
- Regulation of emotions and outcome expectancies, may support GPs in consistently initiating sensitive weight-related conversations.

## Introduction

Overweight, including obesity, remains a major health problem among school-aged children and adolescents. In 2024, 14.8% of Dutch children aged 4-20 years were overweight, 3.5% of whom suffered from obesity (1). Children who are overweight are more likely to remain overweight into adulthood (2), increasing their risk of cardiometabolic diseases (e.g. hypertension and type 2 diabetes) and premature death, and experience more psychosocial issues than peers with a healthy weight status (3). This highlights an urgent need for effective interventions to reduce overweight in children.

In the Netherlands, Youth Health Care assessments become infrequent after age four, with weight and height measured only at ages five and ten (4). Since children visit their general practitioner (GP) for various complaints unrelated to overweight, these consultations can offer an important opportunity to recognize and manage childhood overweight beyond the infrequent Youth Health Care assessments (5). Indeed, the guideline of the Dutch College of GPs (in Dutch: Nederlands Huisartsen Genootschap (NHG)) regarding Obesity advises GPs to identify overweight in children, irrespective of the consultation reason, and suggests a proactive role in the early recognition and management of childhood overweight (6).

Despite this guideline recommendation, GPs infrequently initiate conversations about childhood overweight due to persistent barriers (5). These barriers include practical challenges, such as time constraints and uncertainty regarding referral or treatment options (5, 7), as well as internal processes, like fear of disrupting the doctor–patient relationship or perceiving negative reactions from caregivers (5, 7-9). Internal processes refer to the emotional, cognitive, and decision-making mechanisms that shape a GP’s internal experience and clinical responses. Facilitators include objective assessment tools (e.g., BMI or growth charts), overweight-related presenting complaints, and a trusting relationship with the child and caregivers (7, 8).

Despite increasing insight into the determinants influencing GPs’ initiation of weight-related conversations, the quality of the available evidence remains limited. Existing research largely lacks a theory-driven approach and rely solely on qualitative methods (8, 9), limiting identification of the most influential and modifiable determinants for the development of targeted interventions. Finally, barriers related to internal processes have been minimally explored, with studies focusing on the presence of emotions rather than how GPs regulate them, which may offer opportunities to enhance initiation. For example, adopting an accepting and non-reactive mindset, known as equanimity, has been shown to be beneficial in maintaining emotional balance and psychological distance in challenging situations, including healthcare settings (10).

This study therefore aims to identify the behavioural determinants most relevant and amenable to change for GPs’ initiation of conversations about childhood overweight, with a particular focus on emotional responses and equanimity and their relation to GPs’ intention to initiate such conversations. Our study uses a broad, integrative model of cognitive, emotional, social, and organisational determinants, providing a comprehensive basis to identify key targets for supporting GPs’ initiation of weight-related conversations.

## Methods

### Study design, population and procedure

This cross sectional mixed methods survey study in the Netherlands was open to practicing GPs and GPs in training, with no exclusion criteria. The survey was distributed via email to 1) all GPs within the Extramural LUMC Academic Network (ELAN-GP network) (11), 2) the researchers’ professional network, and 3) via LinkedIn. The email and LinkedIn post included a link to the online survey, which opened with an introduction detailing the study’s objectives, procedures, and data management. Respondents could proceed to the survey by providing their informed consent. The survey was available between March and May 2025, took approximately 15 minutes, and could be completed at the respondent’s convenience.

The study was preregistered on AsPredicted (AsPredicted #229520). The nWMO committee reviewed the proposal and provided a declaration of no objection (nWMO approval number: nr 25-3026). Data were collected anonymously and securely stored in Castor. Access was restricted to project members as needed, with personal data handled according to the General Data Protection Regulation (GDPR).

### Survey

The survey comprised four sections: 1) Respondent characteristics; 2) Determinants of Implementation Behaviour Questionnaire (DIBQ) (12); 3) Self-Assessment Manikin (SAM) (13) to assess emotional valence and arousal, and Equanimity Scale-16 (ES-16) (14), and 4) Behavioural outcomes.

#### Section 1: Respondent characteristics

The first section contained 6 close-ended questions, inquiring on demographical variables.

#### Section 2: DIBQ

To systematically investigate barriers and facilitators, we used the Theoretical Domains Framework (TDF) to assess healthcare professionals’ behaviour. This is a widely used evidence-based framework for this purpose, and comprises 14 domains (15) which are grouped under the COM-B model (Capability, Opportunity, Motivation) at the core of the Behaviour Change Wheel (BCW) (16). The BCW provides a systematic approach for linking behavioural determinants to intervention functions and behaviour change techniques. For quantitative assessment, TDF-based instruments such as the DIBQ (12) have been developed.

We assessed the behavioural determinants of initiating weight-related conversations using a modified version of the validated DIBQ (52 items, 7-point Likert scale) (17). To ensure compatibility with GP practice, the wording of seven statements was slightly modified in collaboration with an experienced GP in the research team, and three items were reverse-coded ([R], see Appendix 1). Two modified items were reassigned to different domains, e.g., ‘The coordination of the intervention is well organized’ became ‘In my workplace there is sufficient coordination regarding initiating conversations about childhood overweight,’ shifting from *Socio-political context to Organization*.

Three items were excluded: two focused on respondents’ confidence and behavioural regulation in initiating conversations in response to experienced barriers, rather than on the nature of these barriers, which was the focus of this study, and the third was not applicable to this context. They were replaced with an item addressing referral options, a relevant barrier in this context (9). Furthermore, items assessing multiple core tasks were reduced to a single task-specific item. Appendix 2 shows all modifications.

The survey also included one closed-ended question on guideline familiarity and two open-ended questions in which initiating was straightforward or challenging.

#### Section 3: SAM and ES-16

We assessed the arousal (intensity) and valence (pleasantness) of emotions when initiating conversations using SAM (see Appendix 1), a simple, intuitive alternative to longer emotion questionnaires, allowing respondents to indicate how positive or negative, and how intense, their feelings were (13).

Equanimity was measured using the validated 16-item ES-16, including the subdomains of non-reactivity and experiential acceptance (14). To preserve the original questionnaire, respondents completed this part independently of their role as a GP, since it has not been validated in healthcare settings. The original English version was translated into Dutch using a forward-backward translation procedure (18). Two researchers (CD and RK) independently translated the ES-16 into Dutch and merged their versions, after which two independent translators back-translated it. A native English speaker slightly refined the items. Eight items were reverse-scored ([R], see Appendix 1).

#### Section 4: Behavioural outcomes

This section included three dependent measures assessed on 7-point Likert scales: 1) Anticipated behaviour: the extent to which respondents plan to initiate (‘how often do you plan to …’, never-always), and two measures of the actual initiation assessing 2) Behavioural frequency, measuring how often GPs report to initiate conversations in practice (‘how often do you think you actually …’, never-always); and 3) Perceived success, indicating how successful they consider themselves in doing so (‘how successful do you consider yourself in …’, not successful-extremely successful). An additional open question asked respondents what they considered necessary to initiate conversations more frequently.

### Data analysis

Data were analysed using R. First, descriptive statistics (mean [SD] or median [IQR], as appropriate) were used to summarize behavioural determinants (DIBQ), with low scores indicating greater room for improvement. Second, the relevance of these determinants was assessed using Pearson correlations between each determinant and the primary outcome, anticipated behaviour (19, 20), as prescribed by the Behaviour Change package (21). Finally, these results were combined into the validated potential for change score (PfC), calculated by subtracting the mean item score from the maximum possible score and multiplying this value by the correlation between the item and anticipated behaviour (20, 21). This relatively novel approach, combing each determinant’s room for improvement with its association with anticipated behaviour, provides a systematic method to prioritize determinants and maximize meaningful behaviour change in quantitative studies.

Behavioural determinants were reported according to their COM-B classification (16), with *Nature of the Behaviours* reported separately (15). The item assessing the pleasantness of initiation (*Beliefs about Consequences*) was excluded due to the conceptual overlap with the *Emotions and Optimism* domain, and because it did not adequately reflect a belief about consequences. Some determinants were combined into composite scales for correlation analyses (see Appendix 3).

For ES-16 and valence/arousal items, descriptive statistics and Spearman correlations with behavioural outcomes and DIBQ items were calculated. ES-16 subdomains (experiential acceptance and non-reactivity) were analysed using aggregated scores (14). Subgroup differences by professional status, sex, age, and working experience, were examined using Spearman correlations or Wilcoxon tests, as appropriate.

Qualitative data on situations where initiating these conversations was easy or difficult were analysed using content analysis guided by the DIBQ framework (22). Two researchers independently analysed the data (CD and RK).

## Results

### Survey and sample characteristics

In total, 193 people accessed the survey, of whom 85 started it. Among these, 62 respondents finished up until section 2 (DIBQ); 57 respondents completed the entire survey. For the analyses, only respondents who completed the full DIBQ were included.

The median age of respondents was 37 years (IQR = 16), and median length of working experience as qualified GP was 9 years (IQR = 15) and for GPs in training 2 (IQR = 2). The majority of respondents were female (*n* = 46, 74.2%); 40.3% of the respondents were GPs in training (*n* = 25). More than half reported being not or partly aware of the guideline of the Dutch College of GPs regarding Obesity (*n* = 34, 54.8%) (Figure 1).

**Figure 1.**
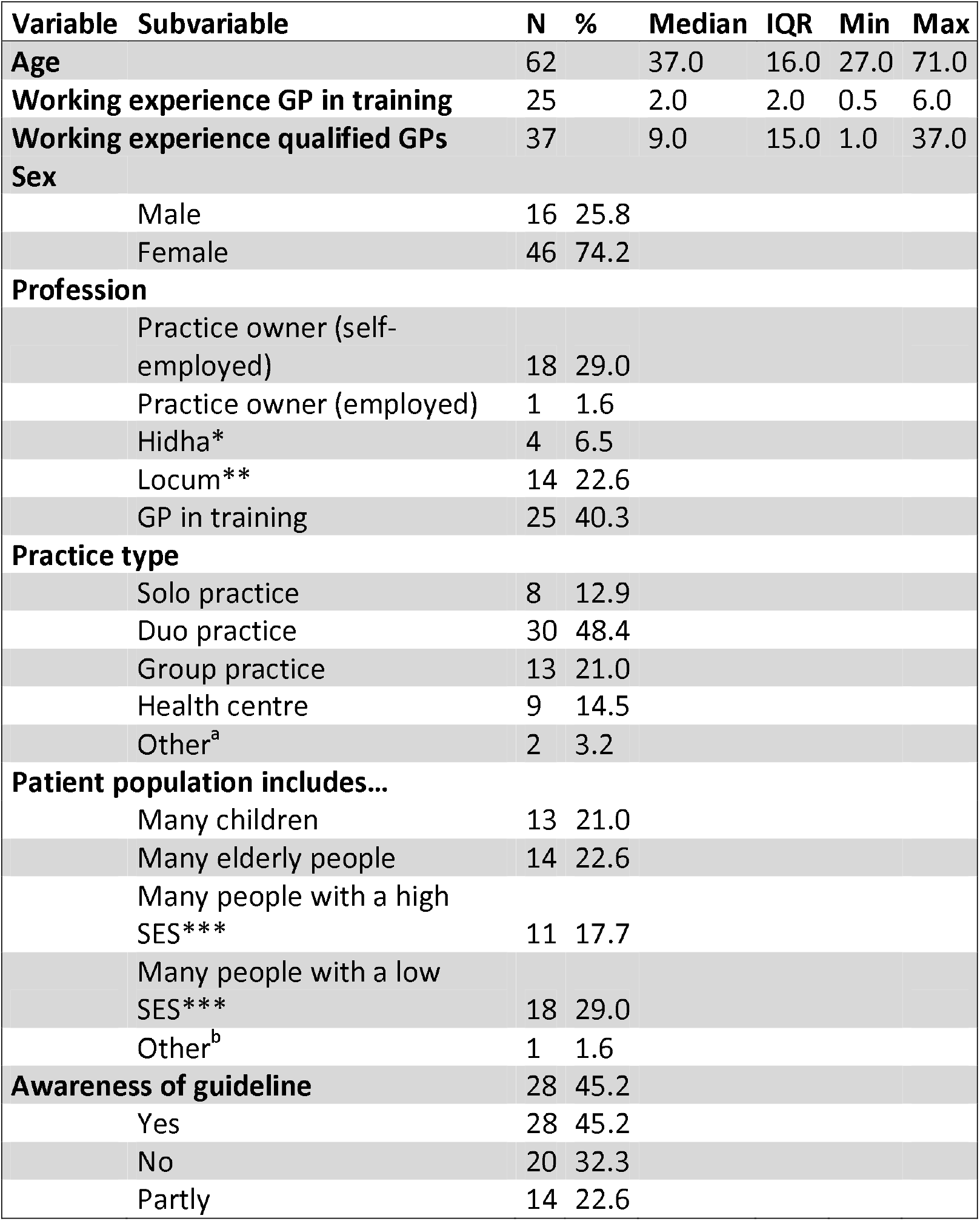
Demographics (n=62) * General Practitioner employed by a General Practitioner; ** Temporary substitute GP; *** Socioeconomic status. a = a GP at the out-of-hours clinic, and one GP in Central Organization for Asylum seekers. b= multicultural: with a large population of Hindustani, Moroccan, and Turkish backgrounds.

### Behavioural outcomes

Among the 57 respondents who completed this section, 53% anticipated initiating conversations about childhood overweight often or always in the future (median = 5.0, IQR = 2.0), compared with 33% reporting rarely or never anticipated doing so. When reflecting on their current behaviour; more than half of the respondents (58%) stated to initiate conversations infrequently (median = 3.0, IQR = 3.0), and 46% of respondents perceived their current success in doing so as low (median = 4.0, IQR = 1.0) (Figure 2).

**Figure 2.**
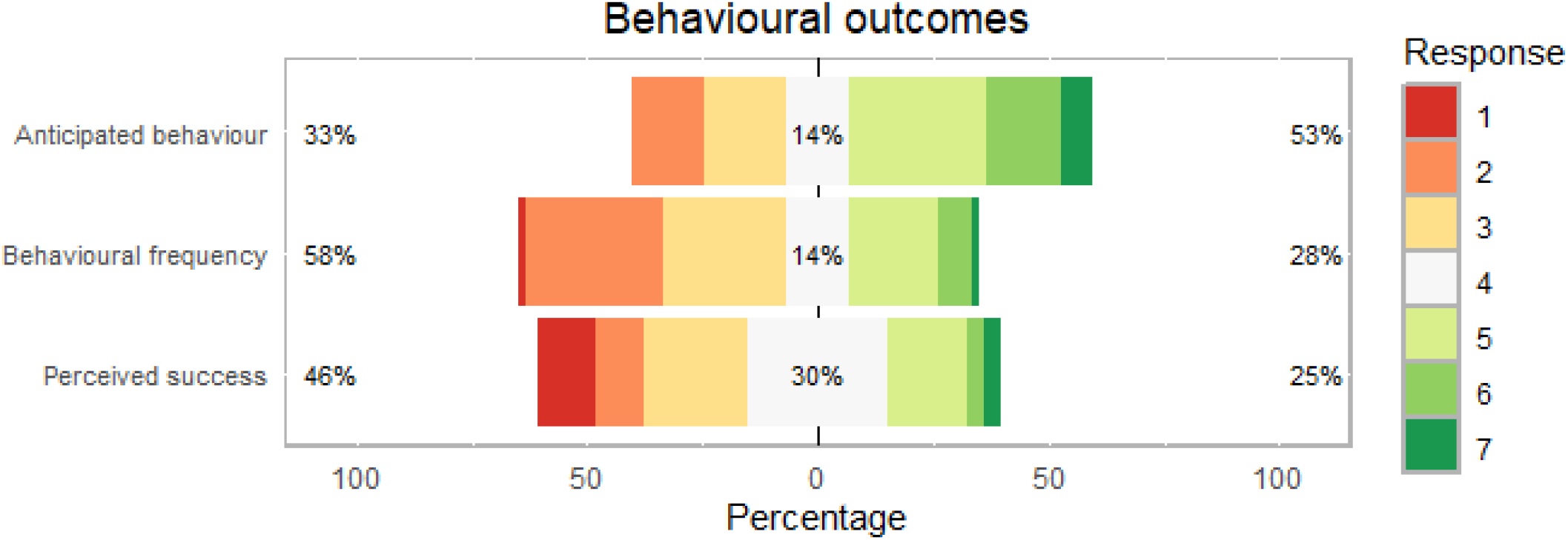
Responses (n = 57) on the items regarding behavioural outcomes on a 7-point scale (for anticipated behaviour and behavioural frequency: 1⍰= never, 7⍰=always, for perceived success: 1= not successful, 7= extremely successful). White indicates neutral (score 4). Left percentages reflect scores 1–3 (low); right percentages reflect scores 5–7 (high).

Appendix 4 presents a subgroup analysis comparing GPs in training with fully qualified GPs. The results show that perceived success was positively associated with age (*r* = 0.338, *p* = 0.010), and that qualified GPs reported higher levels of perceived success than GP trainees (*r* = 0.353, *p* = 0.008).

### DIBQ

#### Descriptive statistics: room for improvement

##### Capability

In total, 62 respondents completed section two of the survey comprising the DIBQ. A majority of respondents (66%) agreed they had sufficient skills (median = 5.0, IQR = 2.0) and 61% agreed to having sufficient knowledge (median = 5.0, IQR = 2.0) to initiate conversations. However, over one third of respondents reported difficulties remembering what to do to initiate conversations, and only 6% of respondents used planning and checking strategies to regulate their behaviour (*Behavioural regulation*) (Figure 3).

**Figure 3.**
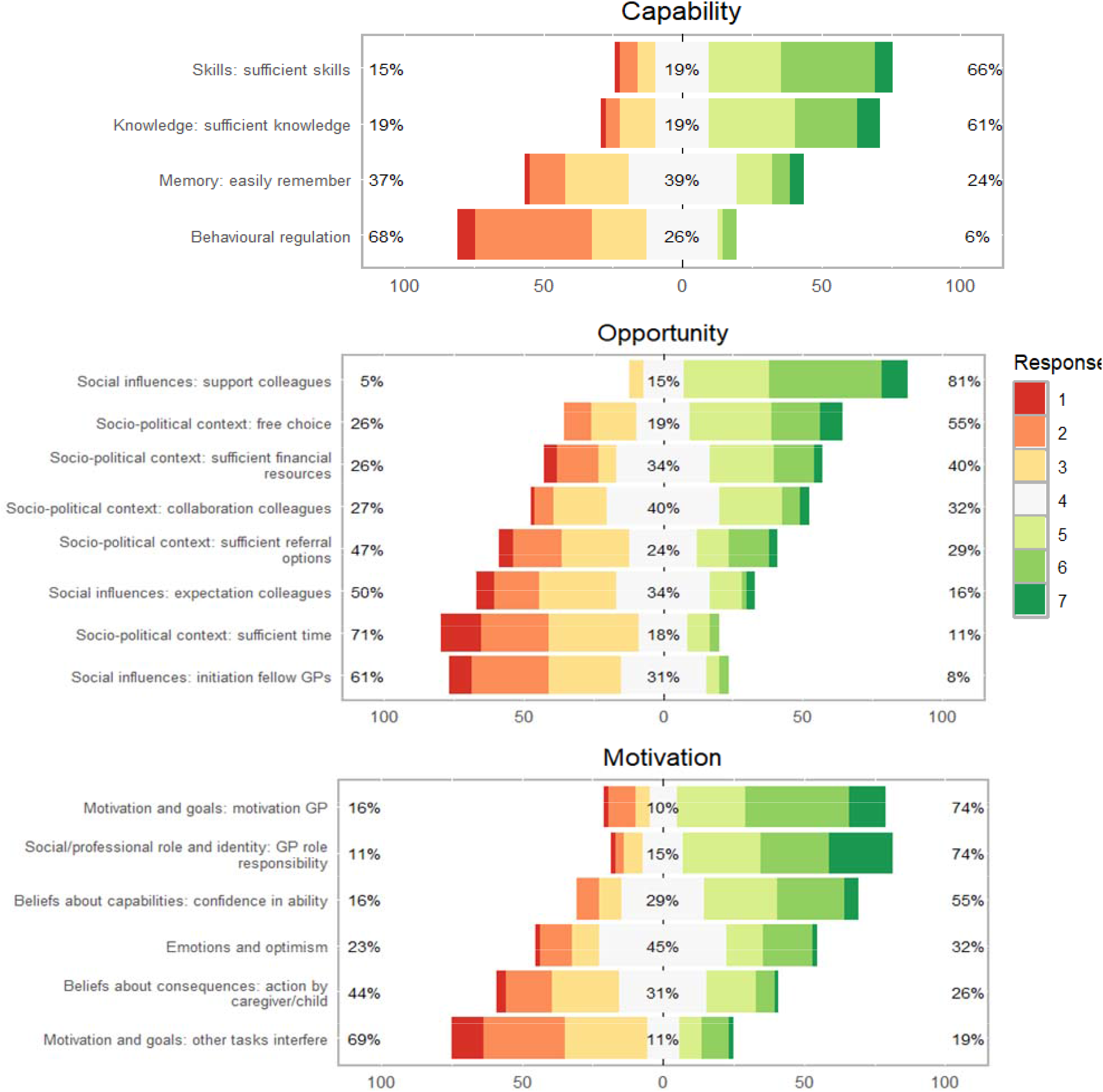
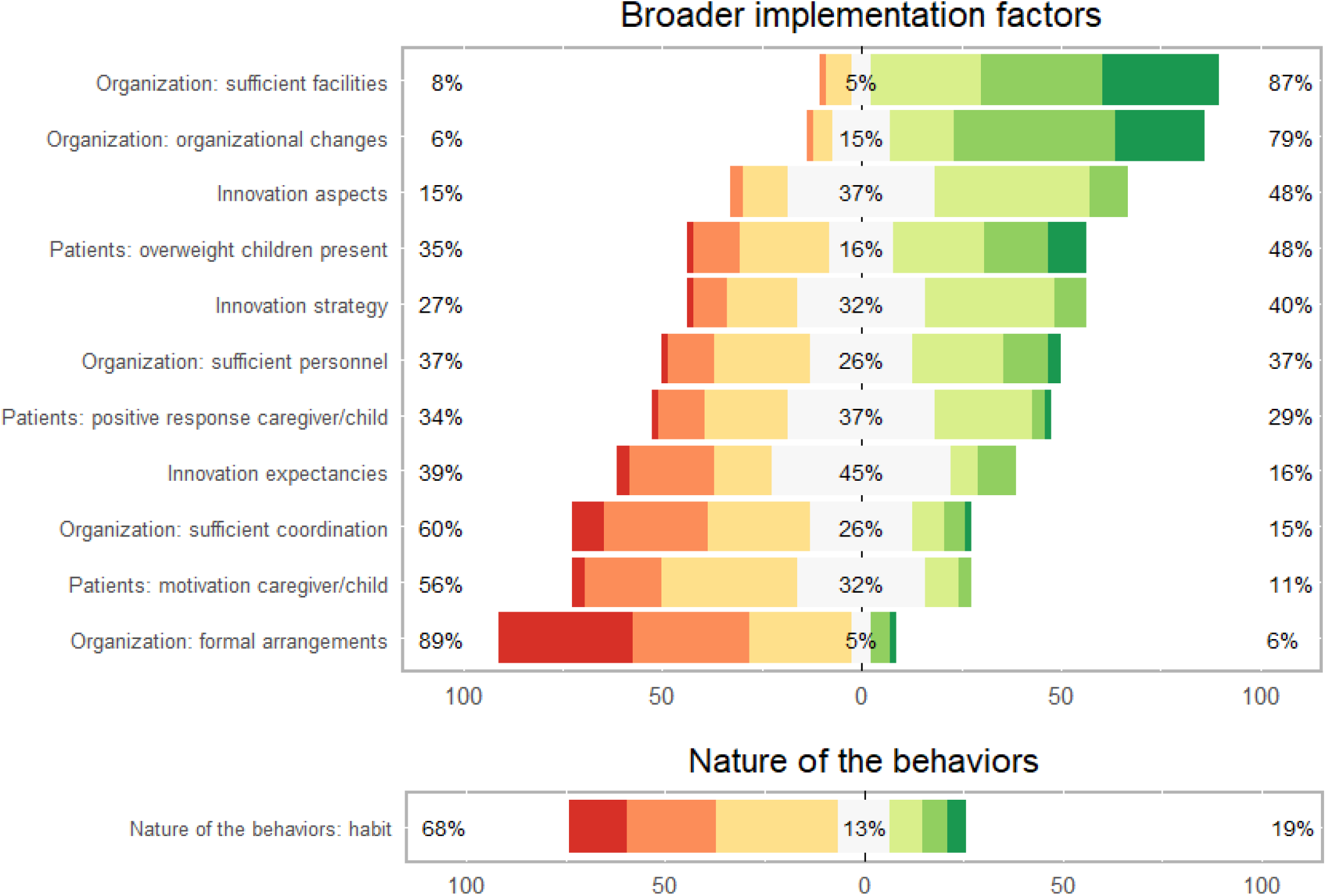
Responses (n = 62) to the DIBQ items, grouped by COM-B classification, on a 7-point scale (1 = totally disagree, 7 = totally agree). White indicates neutral responses (score 4). Percentages on the left reflect scores 1–3; percentages on the right reflect scores 5–7.

##### Opportunity

Most respondents (81%) agreed that they could count on sufficient support from colleagues. In contrast, only 16% agreed that initiating conversations was expected of them and 8% reported that fellow GPs initiate these conversations. Practical barriers were also reported: only 11% indicated having sufficient time, and almost half of the respondents (47%) indicated that they do not have adequate referral options (Figure 3).

##### Motivation

Most respondents (74%) felt motivated and responsible as a GP to initiate conversations, and over half of the respondents (55%) reported feeling confident in their ability to do so (median = 5.0, IQR = 2.0); 16% stated feeling not (entirely) confident. Most respondents responded neutral (31%) or negatively (44%) when asked whether initiating conversations would lead to action (*Beliefs about consequences*). Moreover, 69% indicated that other tasks interfered, and 32% of respondents reported feeling good when initiating conversations, while almost half of respondents (45%) chose a neutral response, indicating they neither felt good nor bad (Figure 3).

##### Broader implementation factors

Most respondents reported having sufficient facilities to initiate conversations (87%). However, 89% reported that no formal arrangements were in place, and 60% indicated insufficient coordination. Barriers related to internal processes were also noted: 16% reported that the effects of initiating conversations were clearly visible and that initiating conversations gives them benefits (*Innovation expectancies*). Additionally, 56% of respondents disagreed that children and caregivers are motivated to take action on weight (Figure 3).

##### Nature of the behaviours

The majority of the respondents (68%) reported that initiating conversations about childhood overweight was not a habitual behaviour (median = 3.0, IQR = 2.0; Figure 3).

##### Correlation analysis: relevance

The strongest correlations with anticipated behaviour were found for the item referring to the motivation of the GP (*r* = 0.62, *p* < 0.001), innovation aspects (*r* = 0.59, *p* < 0.001), and the item referring to initiating conversations about childhood overweight as a habitual action (*Nature of the behaviours*) (*r* = 0.58, *p* < 0.001). These results indicate that higher motivation, perceiving the advice as clear, compatible with routine practice, and applicable, and greater automaticity of this behaviour were associated with more frequent plans to initiate conversations in the future (see Figure 4 and Appendix 5).

**Figure 4.**
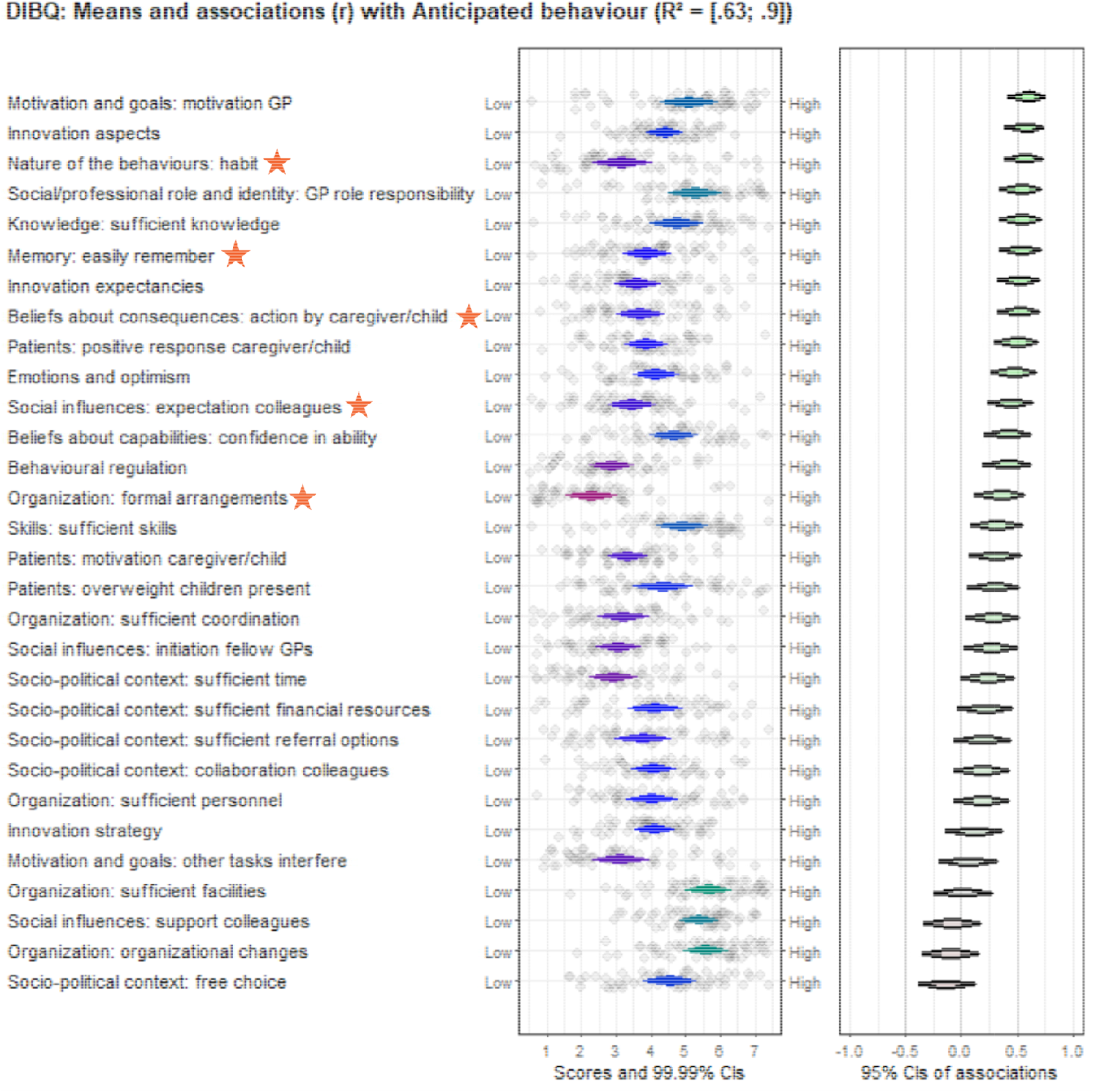
(n = 57) Means (left panel) and associations (right panel) of all determinants with the main outcome anticipated behaviour. Items denoted by a red star indicate the determinants with the highest potential for change scores.

##### Potential for change score (PfC): combining room for improvement with relevance

The highest PfC was observed for the item referring to initiating conversations about childhood overweight as a habitual action (*Nature of the behaviours*, PfC = 2.25). This was driven by both a relatively low mean, indicating considerable room for improvement as the behaviour is not yet habitual, and a strong correlation with anticipated behaviour (*r* = 0.58, *p* < 0.001). Other determinants showing high potential for change included the belief that initiating a conversation prompts caregivers and the child to reflect on the weight and take action (*Beliefs about consequences*, PfC =1.77); the ability to easily recall what needs to be done (*Memory*, PfC =1.73); the presence of formal arrangements within the workplace (*Organization: formal arrangements*, PfC = 1.72); and the expectation from colleagues to engage in such discussions (*Social influences: expectation colleagues*, PfC = 1.64) (Figure 4).

### Equanimity (ES-16)

#### Descriptive statistics

Respondents (*n* = 57) demonstrated, on average, high levels of equanimity on the ES-16 (figure 5) compared to the original validation study (14). Among the two subdomains, respondents scored highest on non-reactivity; only three of the eight items received a score of 1 or 2, and one item had no scores below 3 at all (neutral).

**Figure 5.**
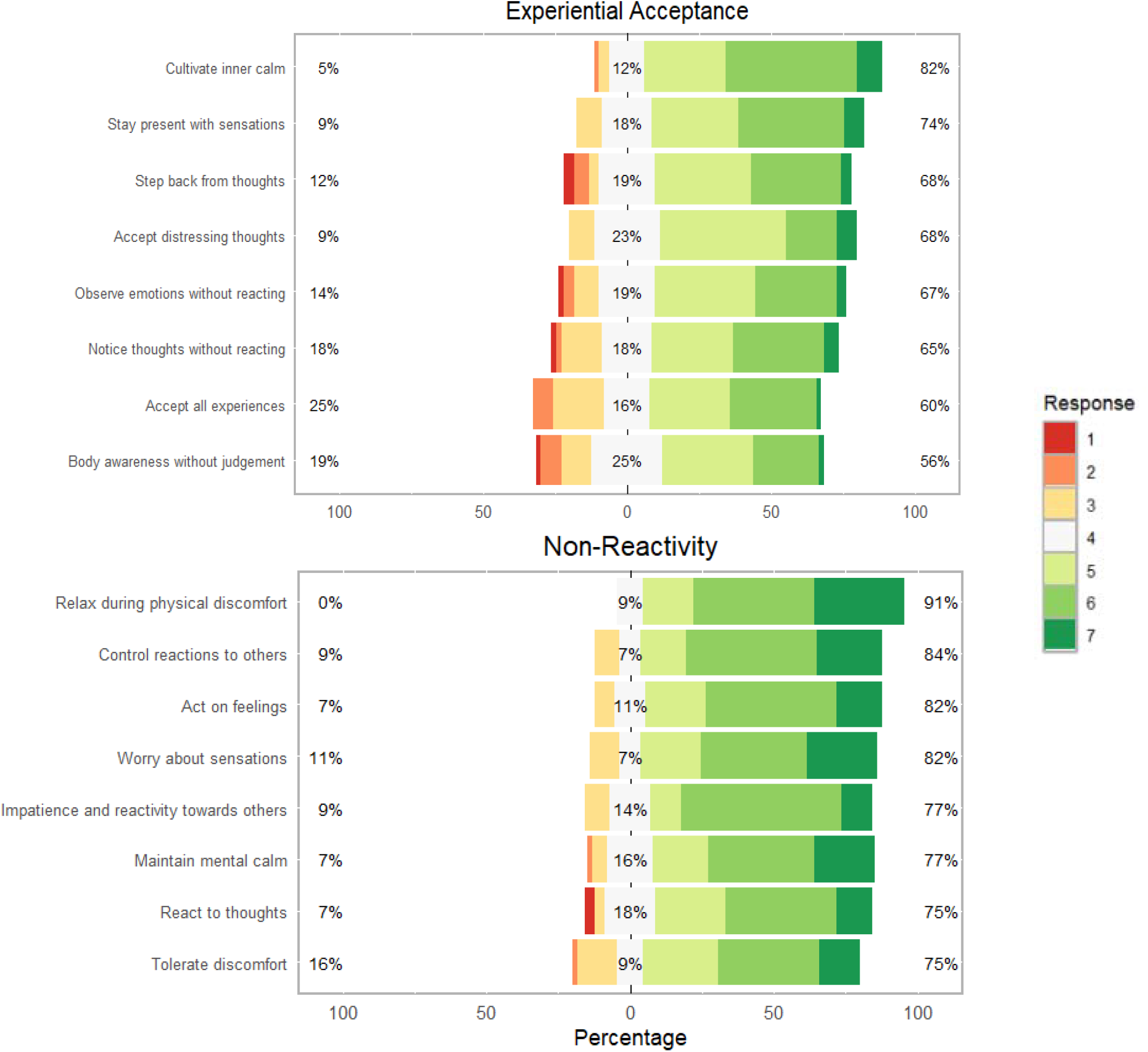
Responses (n = 57) on the ES-16 items (1 = totally disagree, 7 = totally agree). White indicates neutral responses (score = 4). Left percentages represent scores 1–3; right percentages represent scores 5–7. Items are grouped into composite scales (Experiential Acceptance and Non-Reactivity) and renamed for clarity.

The level of equanimity did not differ between GPs in training and qualified GPs, nor between males and females, and was independent of age and years of working experience (see Appendix 4).

#### Correlation analysis

ES-16 scores showed no significant associations with behavioural outcomes, with low correlations across individual items and aggregated subdomains (Appendix 5).

### Valence and arousal (SAM)

#### Descriptive statistics

As illustrated in Figure 6, valence scores, when initiating conversations, were predominantly around the midpoint of the 1-to-9 scale (4 to 5), indicating a generally neutral tendency in respondents’ emotions. Arousal scores were primarily in the range of 3 to 5, indicating that respondents generally experienced moderate emotional activation in this context. A subset of respondents reported more pronounced emotional reactions: 10.2% scored 7 to 9 on arousal, 15.3% scored 1 to 3 on valence indicating that they perceived initiating conversations as negative, and 11.9% scored 7 to 9 on valence, signalling positive emotions.

**Figure 6.**
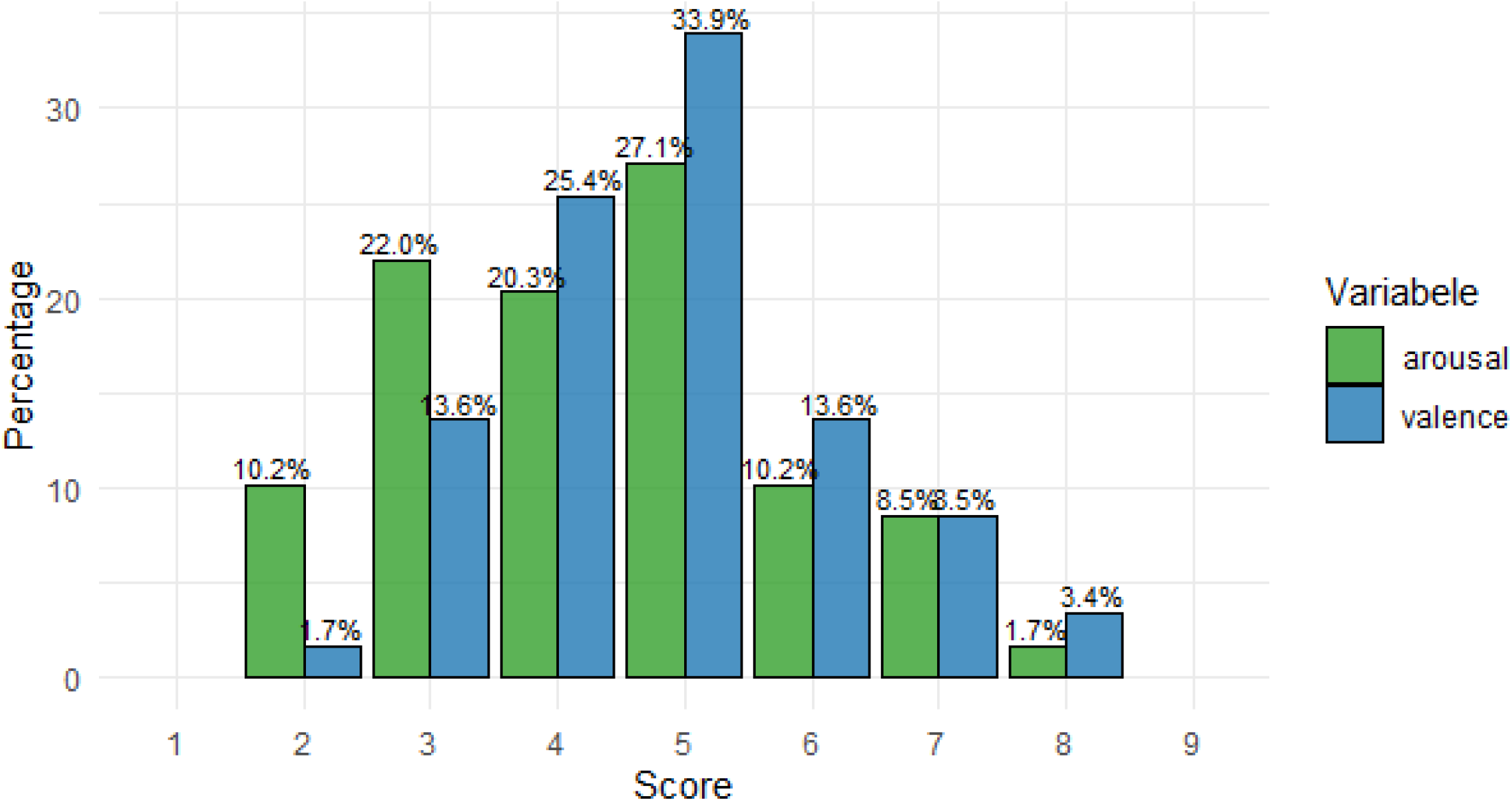
(n = 59) Histogram showing respondents’ rating of valence and arousal when initiating conversations about childhood overweight.

#### Correlation analysis

Valence was positively correlated with behavioural outcomes (anticipated behaviour: *r* = 0.45; frequency: *r* = 0.52; perceived success: *r* = 0.53; all *p* < 0.001), indicating that respondents experiencing more positive emotions were more likely to initiate conversations and generally perceived themselves more successful. In contrast to valence, arousal showed no significant correlations (Appendix 5).

Older respondents scored more positive on valence (*r* = 0.33, *p* = 0.011). Additionally, GPs in training scored more negative compared to fully qualified GPs (*r* = 0.293, *p* = 0.024). Neither working experience nor sex was associated with valence, and arousal was not influenced by any characteristics (Appendix 4).

### Open-ended questions

#### Barriers, Facilitators, and Needs

Figure 7 illustrates the most frequently reported scenarios in which initiating conversations about childhood overweight was perceived as either easy or difficult. Initiating weight-related conversations was perceived as more difficult during consultations addressing unrelated health concerns and easier when the presenting issue was directly linked to weight (e.g., musculoskeletal complaints). Key needs included additional training or tools to initiate the conversation (e.g., specific opening sentences), more consultation time, and greater referral options. Appendix 6 provides an overview of all facilitating and hindering situations, and reported needs.

**Figure 7.**
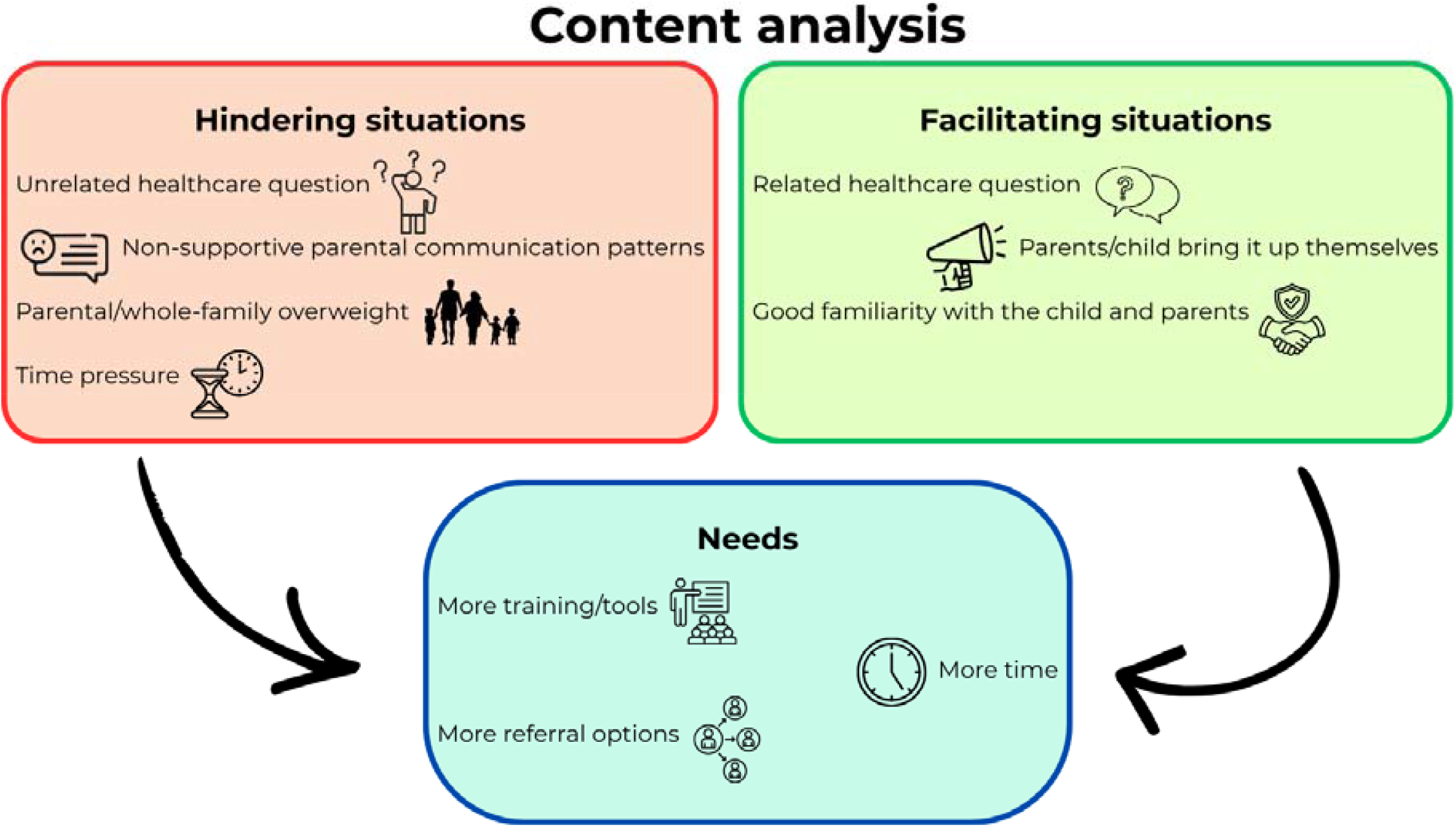
Most commonly reported situations facilitating or hindering the initiation of conversations about childhood overweight, and the most frequently reported needs to support more frequent conversations in general practice.

## Discussion

### Main Findings

This cross-sectional mixed-methods survey study identified behavioural determinants influencing GPs’ initiation of childhood overweight conversations and their potential for change, and furthermore explored how emotional responses and equanimity relate to intention to initiate. While most GPs overall felt relatively knowledgeable, skilled, and motivated, anticipated future and self-reported current behaviour indicated substantial room for improvement. Among the key determinants, practical barriers (e.g. time constraints) and concerns about outcome expectancies (e.g. perceived low child/caregiver motivation) were frequently reported. Making initiation of conversations habitual represented the greatest potential for change, alongside enhancing beliefs that conversations prompt action, improving recall of what to do, and providing supportive formal arrangements within the practice, and a social norm amongst colleague to engage in these discussions. Equanimity was generally high, and only emotional valence (pleasantness), and not arousal (intensity) or equanimity, was correlated with anticipated behaviour of initiating conversations in the future.

### Comparison With Existing Literature and Implications for Future Research

Habit formation, not previously highlighted in this context (8, 9), showed the highest potential for change. This aligns with behavioural science, identifying habit as a key determinant for behaviour change (23), and with the observation that much clinical practice is habitual (24). Habits form through repeated behaviour in stable contexts, creating situation-behaviour associations such that encountering the situation eventually triggers the behaviour automatically, reducing reliance on cognitive resources or motivation (25). In general practice, where time pressure and competing demands are common, habit formation may therefore help ensure that important behaviours are performed reliably. The ability to recall required actions (*Memory*) also showed high potential for change. Memory is particularly important before habit formation, when behaviours still rely on conscious recall, but its role diminishes as behaviour becomes more automatic (26). Similarly, positive outcome expectancies (e.g. that initiation prompts action) support motivation required to engage in and repeat the behaviour initially (27), whereas negative expectancies have been identified as barriers to discussing weight among obese adults (28).

Previous studies identified a lack of social and organisational support as barriers to initiation (8, 9). In line with this, social and organisational factors in the present study showed high potential for change, with colleague expectations and formal arrangements promoting initiating weight-related conversations. These findings align with literature showing that social norm feedback, highlighting peers’ behaviour and expectations, can improve guideline adherence (29).

Overall, our findings align with behavioural science literature, suggesting that behaviour change depends less on knowledge and more on translating knowledge and motivation into action by addressing capability (e.g., memory), motivation (outcome expectancies), opportunity (supportive social and organisational environment), and, most importantly, habit formation. This underscores the value of using a broad, integrative behaviour change model, as applied in this study.

As habit emerged as the determinant with the greatest potential for change, our findings suggest a shift in focus from motivation deficits towards the importance of shaping a context that elicits habitual action. Building on the COM-B model, interventions can support habit formation by identifying practical cues and the most suitable moments in the consultation to initiate conversations (16). By planning in advance how and when to initiate, linking situational cues to specific actions (30), and using memory aids to facilitate recall and repetition, initiating can become more habitual over time. Future research could examine how these habits can be built during GP training and maintained through continuous education.

Children and caregivers being unmotivated to change have been reported as negative outcome expectancies that hinder initiating conversations (8), aligning with our finding that 56% of respondents disagreed that they are motivated to change after initiation. However, it remains unclear whether these expectancies reflect actual caregiver responses or rather assumptions. Future research should examine caregivers’ true motivation, and potential subgroup differences. Even if behaviour change occurs in only a minority of cases, it may still be meaningful given the scale and long-term impact of childhood overweight. Identifying successful cases could help recalibrate expectations and guide targeted strategies.

Additionally, stimulation by the social and organizational context is key to promoting conversations about overweight. Social influences, such as making salient that initiating conversations is a norm amongst colleagues, and organizational support, such as formal arrangements on weighing children, can promote initiation (29). Given that most respondents already felt motivated and responsible to initiate conversations, future research could explore how these positive norms can be made more visible and strengthened.

In addition to standard DIBQ determinants, we examined the role of emotions and emotion regulation, particularly equanimity. Although prior research has offered limited insight into these dynamics, evidence suggests that such conversations are highly sensitive and evoke fear or discomfort (8, 9). Our findings complement this: a small proportion of GPs reported negative valence, and 10.2% of GPs reported substantial arousal. This suggest that these conversations, although considered routine, can be emotionally challenging, indicating room for improvement in regulation of emotions during such interactions. Equanimity, reflecting a broader mindset of emotional stability and non-reactivity, was generally high compared with the original ES-16 validation study, suggesting limited room for improvement (14). Whether this signals that other emotion regulation strategies yield more potential is to be determined as equanimity was measured as a general trait. Therefore scores may reflect overall tendencies rather than situation-specific regulation, which may be markedly different. Future qualitative research could clarify the role of emotions, as well as the efficacy of different emotion regulation approaches, thereby informing the development of targeted interventions.

### Strengths and Limitations

A major strength of this study was using the DIBQ combined with a potential for change analysis, identifying and prioritising determinants most amenable to intervention and complementing previous qualitative research to inform future interventions. Our focus on valence and arousal, as well as equanimity, acknowledges that initiating weight-related conversations can be emotionally challenging and opens up new insights to target other, emotion focused strategies to promote guideline adherence. Including both qualified GPs and trainees enriched the analysis, and focusing specifically on the initiation of conversations offered clearer insights into this critical behavioural threshold.

Potential limitations include self-selection bias, as more motivated GPs may have been more likely to participate, limiting generalizability, and the use of self-report measures, which are susceptible to social desirability and recall bias. Equanimity was assessed as a general trait rather than context specific, limiting its relevance to situation-specific emotional regulation in general practice.

## Conclusion

This study shows that, although GPs overall report intending to initiate conversations about childhood overweight, their behaviour is constrained by a lack of habituation, negative outcome expectancies, failing to remember to act, and a lack of social and organizational support. Moreover, initiating conversations was found to elicit substantial emotion arousal among some GPs, which requires adequate regulation.

Habit formation had the highest potential for change, highlighting the importance of embedding discussions into routine care through repetition in stable contexts. To support habit formation, GPs need assistance to remember to act in busy consultations, positive social norms and formal supportive organizational arrangements. Positive outcome expectancies are also crucial, but promoting them depends on whether GPs’ negative expectancies reflect reality. The emotional sensitivity of these conversations underscores the relevance of awareness and regulation of GPs’ emotions and outcome expectancies. While equanimity was generally high, its role in initiation remains unclear due to a ceiling effect resulting from limited variance. Overall, supporting routine initiation likely requires interventions targeting habit formation, outcome expectancies, memory, and social and organizational influences, alongside further research on how GPs experience and regulate emotions.

## Data Availability

All data produced in the present study are available upon reasonable request to the authors.

## Acknowledgements

We would like to sincerely thank all the GPs who completed the survey for their time and valuable contributions to this study, and dr. L.C. van Gestel for his support with the analyses in R.

