## Appendix 1 for "Determinants of General Practitioners’ Initiation of Conversations about Childhood Overweight: A mixed-methods study"

### Appendix 1: Complete survey

#### Section 1: Respondent characteristics

| Question | Answer options | Type of question |
| --- | --- | --- |
| 1.1 What is your year of birth? | Text | Number (numeric input) |
| 1.2 What is your gender? | Male | Multiple choice (radio buttons) |
|  | Female |  |
|  | Other (free text) |  |
|  | Prefer not to say |  |
| 1.3 What is your current position? | Practice owner (independent) | Multiple choice (radio buttons) |
|  | Practice owner (employed) |  |
|  | Hidha (GP employment contract) |  |
|  | Substitute/Locum |  |
|  | GP trainee (AIOS) |  |
|  | Other (free text) |  |
| 1.4 How many years have you been working in this position? | Text | Number (numeric input) |
| 1.5 What type of practice do you work in? | Solo practice | Multiple choice (radio buttons) |
|  | Duo practice |  |
|  | Group practice |  |
|  | Health center |  |
|  | Other (free text) |  |
| 1.6 What best describes your patient population? | Average population group in the Netherlands | Multiple choice (checkboxes) |
|  | Many children |  |
|  | Many elderly |  |
|  | Many people with a high socioeconomic status |  |
|  | Many people with a low socioeconomic status |  |
|  | Other (free text) |  |

#### Section 2: DIBQ

| **Question** | **Answer options** | **Type of question** |
| --- | --- | --- |
| 2.1 Were you already aware of this advice from the NHG guideline? | Yes | Multiple choice (radio buttons) |
|  | No |  |
|  | Partly |  |
| 2.2 Name at least one specific situation in which it is/was easy for you to start the conversation about childhood overweight. | Text | Text |
| 2.3 Name at least one specific situation in which it is/was difficult for you to start the conversation about childhood overweight. | Text | Text |
| 2.4 To what extent do you agree with the following statements? | Strongly disagree – Strongly agree (scale 1–7) | Slider |
|  | **Label in figures** | |
| It is **clear** to me what is expected of me as a GP regarding the advice from the NHG guideline. | Innovation aspects | |
| The aforementioned advice is **well-constructed.** | Innovation aspects | |
| This advice is compatible with how I am accustomed to work. | Innovation aspects | |
| After initiating the conversation about overweight in children, **the effects** are clearly visible to me. | Innovation expectancies | |
| Initiating the conversation about overweight in children gives me **benefits.** | Innovation expectancies | |
| I have sufficient **knowledge** to initiate conversations about overweight with a child (and parents). | Knowledge: sufficient knowledge | |
| I have sufficient skills to initiate conversations about overweight in children. | Skills: sufficient skills | |
| I think that as a GP, it is my job to initiate the conversation about overweight in children. | Social/professional role and identity: GP role responsibility | |
| I am **motivated** to initiate the conversation about overweight in children. | Motivation and goals: motivation GP | |
| After initiating the conversation, parents and the child reflect on the weight and take action. | Beliefs about consequences: action by caregiver/child | |
| I find it **pleasant** to initiate the conversation about overweight in children. * |  | |
| I am confident that I can initiate the conversation about overweight with the child (and parents) | Beliefs about capabilities: confidence in ability | |
| I feel good when I initiate the conversation about overweight in children. | Emotions and optimism | |
| I feel unpleasant when I initiate the conversation about overweight in children. [R] | Emotions and optimism | |
| Other tasks or things I need to do interfere with initiating the conversation about overweight in children. [R] | Motivation and goals: other tasks interfere | |
| I have **clear plans** of how to initiate the conversation about overweight in children. | Behavioural regulation | |
| I have **sufficient referral options** after I initiated the conversation about overweight in children. | Socio-political context: sufficient referral options | |
| I **check regularly** whether I am doing everything related to initiating the conversation about overweight in children. | Behavioral regulation | |
| I can **easily remember** what I need to do to initiate the conversation about overweight in children. | Memory: easily remember | |
| Initiating the conversation about overweight in children is something I have made my own; it has become a **habit** for me. | Nature of the behaviors: habit | |
| I have **sufficient time** to initiate the conversation about overweight in children. | Socio-political context: sufficient time | |
| I have **sufficient financial resources** available (adequate reimbursement for the consultation) to initiate the conversation about overweight with a child (and parents). | Socio-political context: sufficient financial resources | |
| I experience **the collaboration** with colleagues and other involved parties in initiating the conversation about overweight in children as pleasant. | Socio-political context: collaboration colleagues | |
| Where I work, there is **sufficient coordination** regarding the initiation of the conversation about overweight in children. | Organization: sufficient coordination | |
| Where I work, **formal arrangements** are made with regard to the initiation of conversations about overweight in children (where/when/how). | Organization: formal arrangements | |
| Where I work, there are **sufficient facilities** (a room, scale, measuring tape, etc.) to initiate the conversation about overweight with a child (and parents). | Organization: sufficient facilities | |
| Where I work, **organizational changes** (staff/practice changes) interfere with the initiation of conversations about overweight in children. [R] | Organization: organizational changes | |
| All my fellow GPs initiate the conversation about overweight with the child (and parents). | Social influences: initiation fellow GPs | |
| My colleagues **expect** me to initiate the conversation about overweight with a child (and parents). | Social influences: expectation colleagues | |
| I can count on **sufficient support** from my colleagues regarding the initiation of the conversation about overweight in children. | Social influences: support colleagues | |
| Where I work, there are **sufficient overweight children** for whom the subject of 'overweight' is relevant to discuss. | Patients: overweight children present | |
| Children and parents respond **positively** when I initiate the conversation about overweight. | Patients: positive response caregiver/child | |
| Children and parents are **motivated** to take action regarding weight after I initiated the conversation about overweight in children. | Patients: motivation caregiver/child | |
| I would like to have more **information** to be able to initiate the conversation about overweight in children. | Innovation strategy | |
| I would like to have more **training** to be able to initiate the conversation about overweight in children. | Innovation strategy | |
| I would like to have more **assistance** to be able to initiate the conversation about overweight in children. | Innovation strategy | |
| Initiating the conversation about overweight in children is a **free choice** for me. | Socio-political context: free choice | |
| I get **sufficient recognition** for initiating the conversation about overweight in children. | Innovation strategy | |
| The advice offers **all information and materials** that are necessary to initiate the conversation about overweight in children. | Innovation aspects | |
| It is possible **to tailor** the advice so that it can be applied to any overweight child. | Innovation aspects | |
| I get **sufficient financial reimbursement** for initiating the conversation about overweight in children. | Innovation strategy | |
| Where I work, there is **sufficient personnel** to initiate the conversation about overweight in children. | Organization: sufficient personnel | |
| * This item was removed from the questionnaire and not included in data analysis (see methods section paper) | | |

#### Section 3: SAM and ES-16

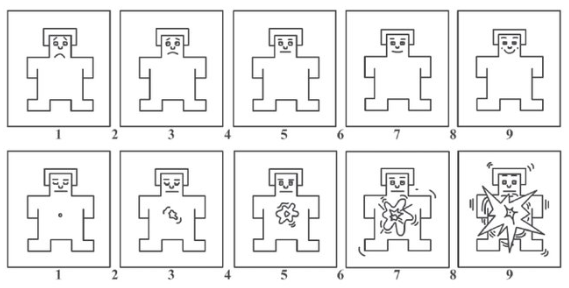

Self-Assessment Manikin (SAM) depicting valence (top) and arousal (bottom)

| **Question** | **Answer options** | **Type of question** |
| --- | --- | --- |
| 3.1 At the moment I have to decide whether to initiate the conversation about overweight with a child (and parents), I experience positive emotions. | Low valence – high valence (scale 1–9) by showing SAM (see above) | Numeric input (text) |
| 3.2 At the moment I have to decide whether to initiate the conversation about overweight with a child (and parents), I feel tense. | Low arousal – high arousal (scale 1–9) by showing SAM (see figure above) | Numeric input (text) |
| 3.3 To what extent do you agree with the following statements? | Strongly disagree – Strongly agree (scale 1–7) | Slider |
|  | **Label in figures** | |
| When I experience distressing thoughts and images, I am able to accept the experience. * | Accept distressing thoughts | |
| I am not able to tolerate discomfort. [R] | Tolerate discomfort | |
| I notice that I need to react to whatever pops into my head. [R] | React to thoughts | |
| When I have a distressing thought or image, I “step back” and am aware of the thought or image without getting taken over by it. * | Step back from thoughts | |
| When I feel physical discomfort, I can’t relax because I am never sure it will pass. [R] | Relax during physical discomfort | |
| I am not able to prevent my reaction when someone is unpleasant. [R] | Control reactions to others | |
| I perceive my feelings and emotions without having to react to them. * | Observe emotions without reacting | |
| I remain present with sensations and feelings even when they are unpleasant. * | Stay present with sensations | |
| I can’t keep my mind calm and clear, especially when I feel upset or physically uncomfortable. [R] | Maintain mental calm | |
| When I notice my feelings, I have to act on them immediately. [R] | Act on feelings | |
| I can pay attention to what is happening in my body without disliking or wanting more of the feeling or sensation. * | Body awareness without judgment | |
| I am impatient and can’t stop my reactivity when faced with other people’s emotions and actions. [R] | Impatience and reactivity towards others | |
| When I have distressing thoughts or images, I am able just to notice them without reacting. * | Notice thoughts without reacting | |
| If I notice an unpleasant body sensation, I tend to worry about it. [R] | Worry about sensations | |
| I approach each experience by trying to accept it, no matter whether it is pleasant or unpleasant. * | Accept all experiences | |
| I endeavor to cultivate calm and peace within me, even when everything appears to be constantly changing. * | Cultivate inner calm | |
| * Items belonging to the Experiential Acceptance subscale | | |

#### Section 4: Behavioral outcomes

| **Question** | **Answer options** | **Type of question** |
| --- | --- | --- |
| If an overweight child visits your consultation for an unrelated complaint… | | |
| 4.1 How often do you intend to initiate the conversation about overweight? | Slider | Never - Always (scale 1-7) |
| 4.2 How often do you think you actually initiate the conversation about overweight in the situation described above? | Slider | Never - Always (scale 1-7) |
| 4.3 How successful do you consider yourself in initiating the conversation about overweight in children? | Slider | Not successful - Extremely successful (scale 1-7) |
| 4.4 What would you need to initiate conversations about childhood overweight more frequently during consultations? | Text | Text |
