## Appendix 3 for "Determinants of General Practitioners’ Initiation of Conversations about Childhood Overweight: A mixed-methods study"

### **Appendix 3 Cronbach’s Alpha and Pearson Correlations for Composite Scales**

| **Domain** | **Items** | **Correlation analysis** | **Cronbach's alfa** |
| --- | --- | --- | --- |
| Behavioural regulation | Behavioural regulation: I have clear plans of how to … | 0.61 (<0.001) |  |
|  | Behavioural regulation: I check regularly whether I am doing everything related to … |  |  |
| Emotions and optimism | Emotions and optimism: I feel good when I … | 0.64 (<0.001) |  |
|  | Emotions and optimism: I feel unpleasant when I … [R] |  |  |
|  |  |  | **Cronbach's alfa** |
| Innovation aspects * | Innovation: It is clear to me what is expected of me as a GP with the advice to … |  | 0.84 (0.76-0.89) |
|  | Innovation: The advice to … is well-constructed |  |  |
|  | Innovation: The advice to … is compatible with how I am accustomed to work |  |  |
|  | Innovation: The advice to … offers all the needed information and materials |  |  |
|  | Innovation: The advice to … can be tailored so that it can be applied to any overweight child |  |  |
|  |  | **Correlation analysis** |  |
| Innovation expectancies * | Innovation: After …, the effects are clearly visible to me | 0.61 (<0.001) |  |
|  | Innovation: … gives me benefits |  |  |
|  |  |  | **Cronbach's alfa** |
| Innovation strategy | Innovation strategy: I would like to have more information to be able to … |  | 0.74 (0.62-0.83) |
|  | Innovation strategy: I would like to have more training to be able to … |  |  |
|  | Innovation strategy: I would like to have more assistance to be able to … |  |  |
|  | Innovation strategy: I get sufficient recognition for … |  |  |
|  | Innovation strategy: I get sufficient financial reimbursement to … |  |  |
|  |  | **Correlation analysis** |  |
| Motivation and goals | Motivation and goals: I am motivated to … | 0.12 (0.37) |  |
|  | Motivation and goals: Other tasks or things I need to do interfere with … [R] |  |  |
|  |  |  | **Cronbach's alfa** |
| Organization | Organization: Where I work, there is sufficient coordination regarding the … |  | 0.57 (0.37-0.72) |
|  | Organization: Where I work, formal arrangements are made with regard to … |  |  |
|  | Organization: Where I work, there are sufficient facilities (a room, scale, measuring tape, etc.) to … | |  |
|  | Organization: Where I work, organizational changes (staff/practice changes) interfere with the … [R] | |  |
|  | Organization: Where I work, there is sufficient personnel to … |  |  |
|  |  |  | **Cronbach's alfa** |
| Patients | Patients: Where I work, there are sufficient overweight children for whom the subject of 'overweight' is relevant to discuss | | 0.61 (0.41-0.75) |
|  | Patients: Children and parents respond positively when I … |  |  |
|  | Patients: Children and parents are motivated to take action regarding weight after I … |  |  |
|  |  |  | **Cronbach's alfa** |
| Social influences | Social influences: My colleagues expect me to ... |  | 0.50 (0.24-0.68) |
|  | Social influences: I can count on sufficient support from my colleagues regarding the … |  |  |
|  | Social influences: All my fellow GPs … |  |  |
|  |  |  | **Cronbach's alfa** |
| Socio-political context | Socio-political context: I have sufficient referral options after I … |  | 0.54 (0.34-0.70) |
|  | Socio-political context: I have sufficient time to … |  |  |
|  | Socio-political context: I have sufficient financial resources to … |  |  |
|  | Socio-political context: I experience the collaboration with colleagues and other involved parties in ... as pleasant | |  |
|  | Socio-political context: … is a free choice for me |  |  |

| Items aggregated into composite scales, justified by Cronbach’s alpha >0.70 (domains with >2 items) or Pearson’s r >0.60 (two-item domains) |
| --- |
| Items not aggregated into composite scales |
| * *Innovation* was further subdivided into *Innovation expectancies,* and *Innovation aspects* to examine the specific role of GPs’ expectancies when initiating conversations. |
