## Appendix 4 for "Determinants of General Practitioners’ Initiation of Conversations about Childhood Overweight: A mixed-methods study"

**Appendix 4 Subgroup analysis**

| *ES-16 total score* |  |  |  |  |  |  |
| --- | --- | --- | --- | --- | --- | --- |
| **GP in training versus Other** | **N** | **Mean** | **SD** | **Median** | **Effect size (r)** | **p-value** |
| GP in training | 23 | 82,217 | 10,436 | 84 | 0,06 | 0,648 |
| Other (all respondents except for GPs in training) | 34 |  |  | 84 |  |  |
| **Male versus female** | **N** | **Mean** | **SD** | **Median** | **Effect size (r)** | **p-value** |
| Male | 15 | 84,400 | 10,528 | 87 | 0,07 | 0,599 |
| Female | 42 | 82,762 | 10,082 | 83,5 |  |  |
| **Variable** | **N** | **Correlation (r)** | **p-value** |  |  |  |
| Age | 57 | 0,118 | 0,382 |  |  |  |
| Working experience | 57 | 0,050 | 0,712 |  |  |  |

***Valence and arousal***

| *Valence* |  |  |  |  |  |  |
| --- | --- | --- | --- | --- | --- | --- |
| **GP in training versus Other** | **N** | **Mean** | **SD** | **Median** | **Effect size (r)** | **p-value** |
| GP in training | 24 | 4,333 | 0,963 | 4 | 0,293 | 0,024 |
| Other (all respondents except for GPs in training) | 35 |  |  | 5 |  |  |
| **Male versus female** | **N** | **Mean** | **SD** | **Median** | **Effect size (r)** | **p-value** |
| Male | 15 | 4,933 | 0,799 | 5 | 0,105 | 0,419 |
| Female | 44 | 4,795 | 1,456 | 5 |  |  |
| **Variable** | **N** | **Correlation (r)** | **p-value** |  |  |  |
| Age | 59 | 0,329 | 0,011 |  |  |  |
| Working experience | 59 | 0,201 | 0,127 |  |  |  |
| *Arousal* |  |  |  |  |  |  |
| **GP in training versus Other** | **N** | **Mean** | **SD** | **Median** | **Effect size (r)** | **p-value** |
| GP in training | 24 | 4,125 | 1,424 | 4 | 0,103 | 0,431 |
| Other (all respondents except for GPs in training) | 35 |  |  | 4 |  |  |
| **Male versus female** | **N** | **Mean** | **SD** | **Median** | **Effect size (r)** | **p-value** |
| Male | 15 | 4,8 | 1,320 | 5 | 0,205 | 0,116 |
| Female | 44 | 4,227 | 1,538 | 4 |  |  |
| **Variable** | **N** | **Correlation (r)** | **p-value** |  |  |  |
| Age | 59 | -0,012 | 0,927 |  |  |  |
| Working experience | 59 | 0,004 | 0,975 |  |  |  |

***Behavioural outcomes***

| *Anticipated behaviour* |  |  |  |  |  |  |
| --- | --- | --- | --- | --- | --- | --- |
| **GP in training versus Other** | **N** | **Mean** | **SD** | **Median** | **Effect size (r)** | **p-value** |
| GP in training | 23 | 4,261 | 1,421 | 5 | 0,04 | 0,765 |
| Other (all respondents except for GPs in training) | 35 |  |  | 5 |  |  |
| **Male versus female** | **N** | **Mean** | **SD** | **Median** | **Effect size (r)** | **p-value** |
| Male | 15 | 4,000 | 1,558 | 4 | 0,124 | 0,35 |
| Female | 42 | 4,452 | 1,501 | 5 |  |  |
| **Variable** | **N** | **Correlation (r)** | **p-value** |  |  |  |
| Age | 57 | -0,015 | 0,909 |  |  |  |
| Working experience | 57 | -0,035 | 0,798 |  |  |  |
| *Behavioural frequency* |  |  |  |  |  |  |
| **GP in training versus Other** | **N** | **Mean** | **SD** | **Median** | **Effect size (r)** | **p-value** |
| GP in training | 23 | 3,174 | 1,267 | 3 | 0,156 | 0,238 |
| Other (all respondents except for GPs in training) | 35 |  |  | 3 |  |  |
| **Male versus female** | **N** | **Mean** | **SD** | **Median** | **Effect size (r)** | **p-value** |
| Male | 15 | 2,933 | 1,223 | 3 | 0,22 | 0,097 |
| Female | 42 | 3,667 | 1,443 | 3 |  |  |
| **Variable** | **N** | **Correlation (r)** | **p-value** |  |  |  |
| Age | 57 | 0,083 | 0,54 |  |  |  |
| Working experience | 57 | 0,045 | 0,74 |  |  |  |
| *Perceived success* |  |  |  |  |  |  |
| **GP in training versus Other** | **N** | **Mean** | **SD** | **Median** | **Effect size (r)** | **p-value** |
| GP in training | 23 | 2,957 | 1,492 | 3 | 0,353 | 0,008 |
| Other (all respondents except for GPs in training) | 35 |  |  | 4 |  |  |
| **Male versus female** | **N** | **Mean** | **SD** | **Median** | **Effect size (r)** | **p-value** |
| Male | 15 | 3,333 | 1,447 | 3 | 0,122 | 0,358 |
| Female | 42 | 3,619 | 1,513 | 4 |  |  |
| **Variable** | **N** | **Correlation (r)** | **p-value** |  |  |  |
| Age | 57 | 0,338 | 0,01 |  |  |  |
| Working experience | 57 | 0,154 | 0,251 |  |  |  |

Significant correlations (p < 0.05)
