## Appendix 6 for "Determinants of General Practitioners’ Initiation of Conversations about Childhood Overweight: A mixed-methods study"

**Appendix 6 Results open-ended questions**

| **Theme** | **Subtheme** | **Number of respondents (n)** |
| --- | --- | --- |
| **Facilitating situations** | | |
| **Related healthcare question** | Related healthcare question according to GP | 44 |
|  | Caregiver attended for obesity; child also appeared obese | 1 |
| **Overweight is reason for consultation** | Caregivers/child bring it up themselves | 12 |
|  | Overweight is the reason for consultation | 2 |
| **Objective measurements available** | GP measures weight/height in all children | 1 |
|  | The child has been weighed | 2 |
| **Doctor–patient relationship (child and caregivers)** | Good familiarity with the child and caregivers | 4 |
|  | Relaxed/good relationship with the child and caregivers | 2 |
| **Acute concern about severity of overweight from GP** | GP has acute concern about the severity of the overweight | 2 |
| **Expected outcome** | Expected caregiver understanding | 1 |
| **Hindering situations** | | |
| **Unrelated healthcare question** | Unrelated healthcare question according to GP | 28 |
|  | The reason for the consultation requires full attention | 2 |
| **Doctor–patient relationship (child and caregivers)** | Poor familiarity with the child and caregivers | 3 |
|  | Difficult relationship with the child and caregivers | 2 |
| **Acute concern about severity of overweight from GP** | Overweight is not severe | 1 |
| **Characteristics caregivers** | Non-supportive caregiver communication patterns | 13 |
|  | Caregivers with a low IQ | 1 |
| **Characteristics child** | Child with low self-esteem/appears insecure | 6 |
|  | Child with psychological complaints | 1 |
| **Characteristics family** | Caregiver and/or whole-family overweight | 7 |
|  | Family issues | 2 |
|  | Cultural/language barrier | 1 |
| **Professional factors** | Time pressure/the reason for the consultation requires full attention | 7 |
|  | Concern about caregivers perceiving the GP as judgmental | 1 |
|  | Uncertainty about whether overweight already has been discussed before | 1 |
| **Needs** | | |
| **Knowledge/skills** | More training | 14 |
|  | More information/tools on how to initiate the conversation | 11 |
|  | More knowledge about support options for caregivers and children | 5 |
| **Motivation and goals** | Greater awareness of the problem/task as a GP | 5 |
|  | Related healthcare question | 2 |
| **Beliefs about capabilities** | More (positive) experience | 4 |
|  | Finding ways to avoid hurting children by addressing the topic | 2 |
| **Beliefs about consequences** | Evidence that initiating the conversation is meaningful | 3 |
| **Socio-political context** | More time | 14 |
|  | More referral options | 9 |
|  | Practice-level/regional agreements about collaboration | 2 |
|  | Cultural change regarding overweight | 1 |
| **Organization** | Practice-level agreements | 3 |
|  | More informational material for caregivers/children | 3 |
|  | Longer tenure within the same practice | 1 |
|  | Standard practice of weighing and measuring every child | 1 |
|  | Visual materials in the consultation room | 1 |
| **Patient** | Trusting relationship with child/caregivers | 1 |
